# The effects of adverse childhood experiences on depression and suicidal behaviors are partially mediated by neuroticism, a forme fruste of major depression

**DOI:** 10.1101/2023.01.31.23285231

**Authors:** Ketsupar Jirakran, Asara Vasupanrajit, Chavit Tunvirachaisakul, Michael Maes

## Abstract

Neuroticism, a personality trait, can predict major depressive disorder (MDD). The current study aims to determine whether a) neuroticism is a feature of the acute state of MDD, including suicidal behaviors (SB); and b) adverse childhood experiences (ACEs) are associated with neuroticism in MDD. This study included 133 participants, 67 normal controls and 66 MDD patients, and assessed the Big 5 Inventory (BFI), ACEs using the ACE Questionnaire, and the phenome of depression using the Hamilton Depression (HAMD) Rating Scale (HAMD), Beck Depression Inventory (BDI), The State-Trait Anxiety Inventory (STAI) and Columbia Suicide Severity Rating Scale (C-SSRS) scores to assess current SB. Neuroticism was significantly higher in MDD than controls, and it explained 64.9% of the variance in the depression phenome (a latent vector extracted from HAMD, BDI, STAI, and current SB scores). The other BFI domains had much less (extraversion, agreeableness) or no effect (openness, conscientiousness). One latent vector could be extracted from the phenome, lifetime dysthymia, lifetime anxiety disorders and neuroticism scores. Neglect (physical and emotional) and abuse (physical, neglect and sexual) account for approximately 30% of the variance in this latent vector. Partial Least Squares analysis showed that the effects of neglect on the phenome were partially mediated by neuroticism, whereas the effects of abuse were completely mediated by neuroticism. Neuroticism (trait) and the MDD phenome (state) are both manifestations of the same latent core, with neuroticism being a less severe manifestation of major depression, which in fact is a multiplicative manifestation of neuroticism.

## Introduction

Globally, mood disorders, such as major depressive disorder (MDD), are among the leading causes of disability (Walker et al., 2015). The effects of depressive symptoms, both on individuals and on society, include diminished quality of life, increased health care costs, increased medical morbidity and mortality (Katon et al., 2003). According to the World Health Organization (WHO), 4.4% of the global population suffers from major depressive disorder (MDD), and approximately 4.4% of Thais suffer from depression (Power, 24 July 2013). The point prevalence of depression is approximately 12.9%, with a one-year prevalence of 7.2% and a lifetime prevalence of 10.8% (Lim et al., 2018).

Depression can occur at any point in life, but it manifests more notably during adolescence, particularly in females (Power, 24 July 2013). Complex interactions between social, psychological, and biological factors are associated with the onset of depressive episodes (Maes and Almulla, 2022). Adverse childhood experiences (ACEs) and negative life events are associated with the onset of depressive episodes (Maes and Almulla, 2022). ACEs, such as emotional neglect and abuse, physical neglect and abuse, and sexual abuse, may also influence the recurrence of illness (ROI) of depressive episodes, lifetime suicidal behaviors including ideation and attempts, and the severity of the phenome of depression, including neurocognitive deficits and anxiety symptoms (Maes et al., 2022b, Maes et al., 2018, Maes and Stoyanov, 2022, Dube et al., 2001, Felitti et al., 1998, Norman et al., 2012). As a result, ACEs affect the course and progression of disease and the lifetime trajectory of patients with depression (Maes et al., 2022a, Maes et al., 2019, Maes et al., 2021).

Importantly, those who have experienced ACEs also have an increased risk of personality disorders and traits, such as neuroticism (Grusnick et al., 2020). For instance, a significant correlation exists between neuroticism and sexual, emotional, and physical abuse, as well as emotional and physical neglect (Roy, 2002). Unlike adverse childhood experiences, negative adult life events have less of an effect on neuroticism (Ogle et al., 2014).

Neuroticism is operationally defined as a personality trait that does not interfere with everyday function but is characterized by a chronic negative emotional state or emotional instability, and increased irritability, anger, sadness, anxiety, worry, self-consciousness, and vulnerability to environmental stressors (Costa and McCrae, 1992; Goldberg, 1993). Moreover, individuals with increased neuroticism experience the environment as unsafe and distressing, and they are frequently self-critical, sensitive to the criticism of others and punishment, they feel personally inadequate, and experience negative affect (Watson et al., 1994; Lamb et al., 2002). Neuroticism is associated with mood and anxiety symptoms in clinical (Malouff et al., 2005) and non-clinical (Saklofske et al., 1995, Jylhä and Isometsä, 2006, Uliaszek et al., 2009) study groups, as well as in cross-sectional studies (Kotov et al., 2010, Griffith et al., 2009). Neuroticism can be measured using the Big Five Inventory (BFI), a rating scale that assesses 5 personality traits, including neuroticism, openness to experience, conscientiousness, extraversion, and agreeableness. However, it is unclear how ACEs and the five BFI personality traits interact, as well as if neuroticism plays a role in mediating how ACEs affect the depressed phenome.

Hence, the aims of the present study are to examine whether: a) ACEs are significantly associated with the 5 BFI personality dimensions in patients with MDD; b) increased neuroticism or lowered levels of the other personality dimensions are associated with the phenome of MDD including suicidal behaviors; and c) the effects of ACEs on the phenome of MDD are mediated via increased neuroticism or lowered openness, conscientiousness, extraversion, or agreeableness.

### Methods and Participants

#### Participants

In this research, we included 67 normal controls and 66 depressed patients. We included Thai-speaking participants, ages 18 - 65 years, both males and females. Patients were recruited as outpatients of the Department of Psychiatry at King Chulalongkorn Memorial Hospital, Bangkok, Thailand. The controls were recruited by word of mouth within the same catchment region, namely Bangkok, from September 2021 to February 2022. The patients were diagnosed as major depressive disorder (MDD) using the DSM-5 criteria (American Psychiatric Association, 2013). The normal controls were staff, and family or friends of staff, and friends of MDD patients. Exclusion criteria for patients and controls are: a) psychiatric axis-1 diagnosis including bipolar disorder, schizophrenia, schizo-affective psychosis, psycho-organic and substance use disorders (SUD) (except tobacco use disorder, TUD; b) axis-2 diagnosis such as antisocial and borderline personality disorder; c) neurological disorders such as stroke, epilepsy, brain tumors and multiple sclerosis, d) major medical illness including (auto)immune disorders, psoriasis, systemic lupus erythematous, inflammatory bowel disease, and rheumatoid arthritis; e) pregnant and lactating women; f) recent surgery; g) infection one month prior to the study; h) subjects treated with immunomodulatory drugs including glucocorticoids and therapeutically doses of antioxidants and omega-3; and i) frequent use of pain killers. In addition, controls were excluded when they suffered from MDD (current and lifetime) and DSM-IV anxiety disorders, and showed a positive family history of mood disorders, suicide and substance use disorders.

All subjects were requested to give voluntary, written informed consent prior to their research participation. The study was conducted in accordance with international and Thai ethical standards and privacy laws. The Institutional Review Board of Chulalongkorn University’s Faculty of Medicine in Bangkok, Thailand (#445/63), authorized the research.

#### Clinical measurements

Socio-demographic and clinical data were obtained using a semi-structured questionnaire obtained through interviews with patients and controls. The semi structured interview comprises sex, marital status, employment status, occupational, income, year of education, family history (FHIS) of mood disorders and suicidal behaviors, and FHIS of substance use disorders (SUD), medical history, psychotropic drugs used, and medical history. MDD was diagnosed using DSM-5 criteria and the Mini International Neuropsychiatric Interview (M.I.N.I.) (Kittirattanapaiboon and Khamwongpin, 2005). The latter case definition and criteria were also employed to assess lifetime and current diagnoses of dysthymia, generalized anxiety disorder (GAD), panic disorder (PD), agarophobia, social phobia, obsessive-compulsive disorder (OCD), post-traumatic stress disorder (PTSD), and SUD. We used the DSM-IV concept of anxiety disorders, namely PTSD, GAD, OCD, phobia, agoraphobia, and PD to make the diagnosis of lifetime or current anxiety disorder (Maes et al., 2022a). The severity of depression was measured using the Hamilton Rating Scale for Depression (HAM-D) (Hamilton, 1960) and the Beck Depression Inventory II (BDI-II) (Beck et al., 1996). The latter is a 21-item self-report inventory designed to assess the presence and severity of depressive symptoms and was translated into Thai by Mungpanich (Mungpanich, 2008). The severity of anxiety was assessed using the State-Trait Anxiety Inventory (STAI) state version (Spielberger et al., 1983) in a validated Thai translation (Iamsupasit and Phumivuthisarn, 2005). The DSM-5 criteria were used to make the diagnosis of TUD.

The Big Five Inventory (BFI) (John and Srivastava, 1999) in a Thai translation (Luangsurong, 2016) was used to assess five major personality dimensions, namely neuroticism, openness to experience, conscientiousness, extraversion, and agreeableness. This scale consists of 44 questions rated on a Likert scale scoring from 1 (lowest) to 5 (highest). In the present study, we used the raw scores of the 5 dimensions as well as factors extracted from a combination of these dimensions. ACEs were assessed using the ACE Questionnaire in a Thai translation (Rungmueanporn et al., 2019). This questionnaire consists of 28 items covering the traumatic experiences in childhood in 10 domains, namely: emotional abuse (2 items), physical abuse (2 items), sexual abuse (4 items), emotional neglect (5 items), physical neglect (5 items), domestic violence (4 items), household substance abuse (2 items), mental illness in household (2 items), parental divorce (1 item), and a criminal household member (1 item). The content validity of questionnaire met the standards, with an internal consistency reliability of 0.79 for the abuse domain, 0.82 for the neglect domain and 0.66 for the household dysfunction domain. We used the Columbia Suicide Severity Rating Scale (C-SSRS) (Posner et al., 2011) to assess the intensity of suicidal ideation and suicidal attempts, either lifetime or current. The C-RRRS measures the severity of suicidal ideation (SI) and attempts (SA), intensity, lethality, and frequency, as well as self-injurious behaviors without suicidal intent.

The ASSIST (Alcohol, Smoking and Substance Involvement Screening Test) is a tool to evaluate experiences with alcohol consumption, tobacco (TUD) and substance (SUD) abuse (Assanangkornchai et al., 2014). This tool focuses on the substance used in the last 3 months. Body mass index (BMI) was calculated as weight (kg) divided by height (m) squared. Metabolic Syndrome (MetS) was defined according to the American Heart Association/National Heart, Lung, and Blood Institute 2009 Joint Scientific Statement (Alberti et al., 2009) as the presence of 3 or more of the following components: (1) waist circumference ≥ 90 cm for men, ≥80 cm for women or BMI ≥ 25 kg/m2 ; (2) high triglyceride level: ≥ 150 mg/ dL (≥ 1.69 mmol/L); (3) low HDL cholesterol level: < 40 mg/ dL (< 1.03 mmol/L) for men, < 50 mg/dL (< 1.29 mmol/L) for women; (4) high blood pressure: ≥ 130 mm Hg systolic blood pressure, ≥ 85 mm Hg diastolic blood pressure, or treatment with antihypertensive medication; and (5) high fasting glucose (≥ 100 mg/dL [≥ 5.56 mmol/L]) or a diabetes diagnosis.

#### Statistical analysis

ANOVA was used to compare scale variables among diagnostic groups, while chisquare or Fisher’s Exact Probability Test was employed to compare nominal variables across categories. Correlations between two sets of scale variables were computed using Pearson’s product moment or Spearman’s rank order coefficients, while associations between scale and binary variables were examined using point-biserial correlation coefficients. Results of multiple comparisons or correlations are p-corrected for false discovery rate (FDR). We performed principal component analysis (PCA) to reduce the number of items (e.g., ACE, BFI domains) into one PC score, which then could be used in other statistical analyses. Factorability was checked using the Kaiser-Meyer-Olkin test for sample adequacy, which is considered satisfactory when > 0.5, and the Bartlett’s sphericity test. The first PC is only accepted when the variance explained (VE) is > 50% and all loadings on the first PC are > 0.65. The effects of explanatory variables (ACEs, Big 5 scores) on dependent variables (such as the phenome score) were examined using multiple regression analysis (manual method). In addition, we used a forward stepwise automatic regression method with p-values of 0.05 to-enter and 0.1 to remove to delineate the best predictors of the model. In addition to F statistics (and p values) and total variance (R^2^ or partial eta squared as effect size) explained by the model, we calculated the standardized coefficients with t-statistics and exact p-values for each of the explanatory variables in the final regression models. Collinearity and multicollinearity were analyzed utilizing tolerance (cut-off value <0.25), the variance inflation factor (cut-off value >4), the condition index and variance proportions from the collinearity diagnostics table. The White and modified Breusch-Pagan tests were utilized to confirm the existence of heteroskedasticity. All of the aforementioned tests were two-tailed, and an alpha value of 0.05 was considered statistically significant. We employed the IBM, Windows SPSS version 28.

Partial Least Squares path analysis (SmartPLS) was used to determine the causal relationship between the ACEs (input variables) and the phenome of depression (output variable). All variables were entered either as latent vectors (LVs) derived from their manifestations or as single indicators. When the inner and outer models met predefined quality criteria, such as a) the model fit is < 0.08 in terms of standardized root mean squared residual (SRMR), b) the LVs have a high composite reliability (> 0.7), Cronbach’s alpha (> 0.7), and rho A (> 0.8) values, with an average variance extracted (AVE) > 0.5, and c) all LV loadings are greater than 0.65 at p<0.001, a complete PLS analysis was performed using 5,000 bootstrap samples. We also ran a Confirmatory Tetrad Analysis (CTA) to make sure the LVs were not misclassified as reflective models. Using PLS predict and a tenfold cross validation technique, the model’s prediction performance was tested. We computed pathway coefficients (with exact p values) as well as the total direct and indirect effects and specific indirect effects. An a priori power calculation to estimate the sample size using G*Power 3.1.9.4 showed that to obtain a power of 0.8 in a multiple regression analysis (or PLS analysis) with 5 covariates, the sample size should be at least 70 when using an effect size of 0.2 at p=005 (two-tailed).

### Results

#### Sociodemographic data of patients and controls

**Table 1** shows that there were no significant differences in sex ratio, age, education, BMI, waist circumference, income, TUD, systolic and diastolic blood pressure between controls and patients. Only the employment rate was significantly different between both groups.

**Table 1.** Demographic and clinical data of the Major Depression patients and healthy controls (HC) included in the present study.

| **Variables** | **HC <sup>a</sup>**<br>**(n=67)** | **Major depression <sup>b</sup>**<br>**(n=66)** | **F/X<sup>2</sup>/FEPT** | **df** | **p** |
| --- | --- | --- | --- | --- | --- |
| Sex (Male / Female) | 9 / 58 | 18 / 48 | 3.94 | 1 | 0.055 |
| Age (years) | 37.9 (9.2) | 37.0 (11.5) | 0.263 | 1/131 | 0.609 |
| Education (years) | 14.1 (3.4) | 14.8 (2.8) | 1.72 | 1/131 | 0.193 |
| Employment (No/Yes) | 0/66 | 19/47 | 19.388 | 1 | <0.001 |
| Income (baht/month) | 21500 (7269) | 23770 (19203) | 0.533 | 1/133 | 0.467 |
| BMI (kg/m<sup>2</sup>) | 27.39 (6.11) | 27.85 (9.66) | 0.108 | 1/131 | 0.743 |
| Waist Circumference | 88.8 (14.9) | 89.7 (15.0) | 0.142 | 1/130 | 0.707 |
| Systolic Blood Pressure | 127.9 (15.3) | 129.9 (17.8) | 0.475 | 1/131 | 0.492 |
| Diastolic Blood Pressure | 79.2 (11.3) | 77.9 (12.4) | 0.420 | 1/131 | 0.518 |
| Metabolic syndrome (No/Yes) | 46/21 | 50/16 | 0.84 | 1 | 0.440 |
| Anxiety disorders (lifetime + current) | 67/0 | 48/18 | 21.13 | 1 | <0.001 |
| Dysthymia (lifetime + current) | 67/0 | 42/24 | 29.73 | 1 | <0.001 |
| Tobacco use disorder (No/Yes) | 62/5 | 54/12 | 3.427 | 1 | 0.064 |
| Antidepressants (No/Yes) | - | 8/58 |  |  |  |
| Benzodiazepine (No/Yes) | - | 30/36 |  |  |  |
| Antipsychotic (No/Yes) | - | 52/14 |  |  |  |
| Mood stabilizer (No/Yes) | - | 64/2 |  |  |  |
| Medication Hypertension (No/Yes) | 56/11 | 61/5 | 2.456 | 1 | 0.117 |
| Medication Diabetes (No/Yes) | 61/6 | 61/5 | 0.083 | 1 | 0.773 |
| Medication Dyslipidemia (No/Yes) | 58/9 | 55/11 | 0.272 | 1 | 0.602 |

Results are shown as mean ± SD. F: results of analysis of variance; X<sup>2</sup>: analysis of contingency tables, FFHT: Fisher-Freeman-Halton Exact Test. BMI: body mass index. ASSIST-Tobacco; Alcohol, Smoking and Substance Involvement Screening Test -Tobacco

#### Results of PC analysis

**Table 2** shows the results of PC analyses. We were not able to extract one PC from all ACE items, however, three validated PCs could be constructed. First, we were able to extract a PC from ACE9, ACE10, ACE11, ACE12, ACE13 and ACE15 and labeled this “PC_neglect” (see **Electronic Supplementary File (ESF) Table 1** for explanation of the ACE items). A second validated factor could be extracted from 8 other symptoms, including ACE1, ACE2, ACE3, ACE4, ACE19, ACE20, ACE21 and ACE22, dubbed “PC_abuse”. We were able to extract a third validated PC from ACE5, ACE6, ACE7 and ACE8, and dubbed this “PC_sexabuse”. Other items (including the ACE divorce item) were entered as single indicators in the analysis.

**Table 2.** Results of principal component (PC) analyses

| PC_neglect |  | PC_abuse |  | PC_sexabuse |  | PC_4BFI |  | Phenome |  | TS_phenome |  |
| --- | --- | --- | --- | --- | --- | --- | --- | --- | --- | --- | --- |
| Variables | Loadings | variables | Loadings | Variables | Loadings | Variables | Loadings | Variables | Loadings | Variables | Loadings |
| ACE9 | 0.820 | ACE1 | 0.748 | ACE5 | 0.687 | Extra | 0.787 | HDRS | 0.920 | Neuro | 0.864 |
| ACE10 | 0.864 | ACE2 | 0.748 | ACE6 | 0.862 | Agree | 0.742 | BDI | 0.938 | Dysthymia | 0.736 |
| ACE11 | 0.932 | ACE3 | 0.795 | ACE7 | 0.888 | Consc | 0.756 | STAI | 0.892 | Anxiety | 0.708 |
| ACE12 | 0.909 | ACE4 | 0.739 | ACE8 | 0.787 | Neuro | -0.760 | SB | 0.736 | Phenome | 0.897 |
| ACE13 | 0.854 | ACE19 | 0.773 |  |  |  |  |  |  |  |  |
| ACE15 | 0.776 | ACE20 | 0.824 |  |  |  |  |  |  |  |  |
|  |  | ACE21 | 0.725 |  |  |  |  |  |  |  |  |
|  |  | ACE22 | 0.688 |  |  |  |  |  |  |  |  |
| KMO=0.869 |  | KMO=0.821 |  | KMO=0.729 |  | KMO=0.776 |  | KMO=0.809 |  | KMO=0.716 |  |
| X<sup>2</sup>=679.13 (df=15) |  | X<sup>2</sup>=697.12 (df=28) |  | X<sup>2</sup>=219.48 (df=6) |  | X<sup>2</sup>=117.48 (df=6) |  | X<sup>2</sup>=376.63 (df=6) |  | X<sup>2</sup>=225.66 (df=6) |  |
| p<0.001 |  | p<0.001 |  | p<0.001 |  | p<0.001 |  | p<0.001 |  | p<0.001 |  |
| VE=74.09% |  | VE=57.18% |  | VE=65.58% |  | VE=58.0% |  | VE=76.60% |  | VE=64.87% |  |

ACE : Adverse Childhood Experiences (see Electronic Supplementary File, table 1, for explanation); Extra : Big Five Inventory extraversion dimension; Agree : Big Five Inventory agreeableness dimension; Consc : Big Five Inventory conscientiousness dimension; Neuro : Big Five Inventory neuroticism dimension; HDRS: Hamilton Depression Rating Scale; BDI : Beck Depression Inventory; STAI: Spielberger State and Train Anxiety, State version; SB : Suicidal behaviors; Anxiety: any DSM-5 anxiety disorder, KMO : Kaiser-Meyer-Olkin; VE: variance explained

Table 2 also shows that we were able to extract one PC from 4 BFI dimensions, namely extraversion, agreeableness, consciousness (all positively loaded on the first PC) and neuroticism (negative loaded). Consequently, we used the inverse transformation (to underscore the role of neuroticism) of this PC in the analysis, and we labeled this indicator “PC_4BFI” (which thus reflects the effects of neuroticism and lowered extraversion, agreeableness, and conscientiousness). The fifth BFI dimension (openness to experience) did not load significantly on this first PC (loading of 0.323). Current suicidal ideation (dubbed: SI) was computed as the first PC extracted from 7 C-SSRS items, namely C-SSRS11, C-SSRS12, C-SSRS13, C-SSRS15, C-SSRS16 and C-SSRS17 (Maes et al., 2022) (KMO=0.703, Bartlett’s χ2=982.510, df=15, p<0.001, explained variance=72.05%, all loadings > 0.672). See ESF Table 2 for an explanation of the CSSRS items. The current SA (dubbed SA) score was computed as the first PC extracted from 5 C-SSRS items, namely C-SSRS30, C-SSRS31, C-SSRS32, C-SSRS33 and C-SSRS34 (Maes et al., 2022) (KMO=0.636, Bartlett’s χ2=302.236, df=3, p<0.001, explained variance=81.14%, all loadings > 0.807). Current suicidal behaviors (SB) were conceptualized as the first PC extracted from PC_SI and PC_SA.

We were also able to extract one PC from the HAMD, BDI, STAI and current SB scores, and labeled this first PC “phenome” (see Table 2). Moreover, as shown in Table 2 we were able to extract one PC from neuroticism, lifetime dysthymia, any lifetime anxiety disorders, and the phenome, and dubbed this PC “trait-state-(TS)-Phenome”.

#### ACEs and BFI scores in MDD

**Table 3** shows that the PC_neglect, PC_abuse, PC_sexabuse and ACE_divorce scores were significantly higher in MDD than controls. The frequency of FHIS of mood disorders and SBs, and FHIS of SUD were significantly higher in patients than in controls. The BFI extraversion and agreeableness scores were significantly lower in MDD than in controls, whereas the BFI neuroticism score was significantly higher in MDD. There were no significant differences in openness and conscientiousness scores between both groups. All clinical scores were significantly higher in patients than in controls.

**Table 3.**
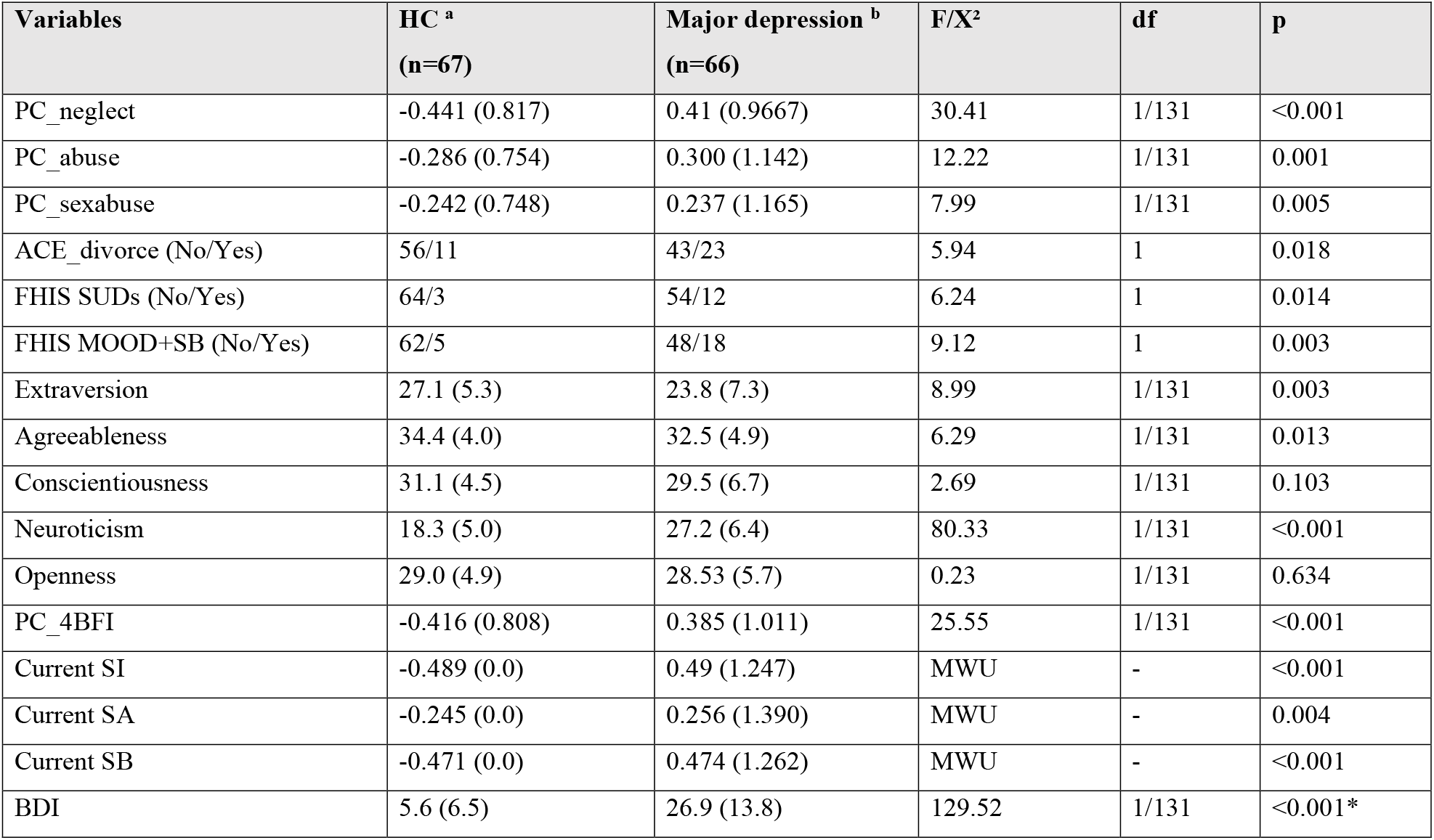
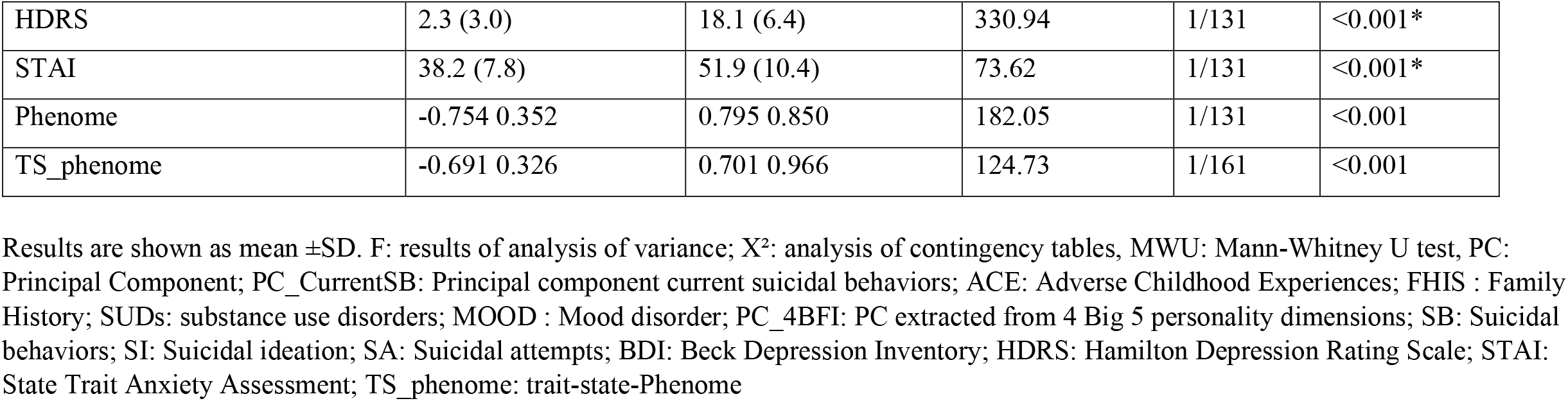
Clinical features of major depressive disorder (MDD) and healthy controls (HC), including adverse childhood experiences (ACE), personality dimensions as assessed with the Big Five Inventory (BFI), assessment of depression and anxiety severity, and lifetime and current suicidal behaviors

#### Intercorrelation matrix

**Table 4** shows the intercorrelations between the different variables measured in our study. In the total study group, PC_neglect was significantly correlated with extraversion, agreeableness, conscientiousness, and neuroticism. In MDD subjects (n=66), PC_neglect was significantly associated with extraversion (r=-0.258, p=0.037). In the total study group, there were significant correlations between PC_abuse and all BFI dimensions, except openness, whereas in MDD patients there were significant associations with agreeableness (r=-0.252, p=0.041), conscientiousness (r=-0.313, p=0.010) and neuroticism (r=0.486, p<0.001). Table 4 shows that in the total study group, PC_sexabuse was strongly associated with neuroticism but also with openness. In subjects with MDD, there was a strong positive association between ACE_sexabuse and openness (r=0.339, p=0.005).

**Table 4.** Intercorrelation matrix.

| Variables | PC_neglect | PC_abuse | PC_sexabuse | PC_currentSB | PC_phenome |
| --- | --- | --- | --- | --- | --- |
| PC_neglect | 1.0 | 0.448 (<0.001) | 0.166 (0.055) | 0.339 (<0.001) | 0.512 (<0.001) |
| PC_abuse | 0.448 (<0.001) | 1 | 0.234 (0.007) | 0.346 (<0.001) | 0.458 (<0.001) |
| PC_sexabuse | 0.133 (0.166) | 0.234 (0.007) | 1 | 0.236 (0.006) | 0.342 (<0.001) |
| Extraversion | -0.301 (<0.001) | -0.281 (0.001) | -0.114 (0.190) | -0.208 (0.016) | -0.381 (<0.001) |
| Agreeableness | -0.285 (<0.001) | -0.287 (<0.001) | 0.038 (0.662) | -0.181 (0.038) | -0.372 (<0.001) |
| Conscientiousness | -0.206 (0.017) | -0.238 (0.006) | -0.169 (0.051) | -0.207 (0.017) | -0.330 (<0.001) |
| Neuroticism | 0.430 (<0.001) | 0.504 (0.001) | 0.349 (<0.001) | 0.458 (<0.001) | 0.837 (<0.001) |
| Openness | -0.021 (0.814) | -0.015 (0.866) | 0.173 (0.047) | 0.091 (0.298) | -0.077 (0.380) |

Table 4 shows that SB was significantly correlated with PC_neglect, PC_abuse, PCsexabuse, and all BFI scores (except openness); the strongest correlation was established with neuroticism. In MDD patients, SB was only correlated with neuroticism (r=0.300, p=0.014). The same table shows that the severity of the phenome was predicted by all PC_ACE scores, and all BFI scores (except openness). There was a very strong association between phenome and neuroticism with r=0.837. In the restricted study group of MDD patients, we found strong associations between phenome and PC_neglect (r=0.319, p=0.005), PC_abuse (r=0.345, p=0.005), extraversion (r=-0.368, p=0.002), agreeableness (r=-0.264, p=0.032), conscientiousness (r=-0.358, p=0.003), and neuroticism (r=0.643, p<0.001). In MDD patients, there were significant point-biserial associations between dysthymia and extraversion (r=-0.258, p=0.037), neuroticism (r=0.295, p=0.016) and significant associations between any anxiety disorder and PC_abuse (r=0.359, p=0.003), extraversion (r=-0.263, p=0.033), neuroticism (r=0.401, p<0.001), and openness (r=0.243, p=0.049).

#### Results of multiple regression analysis

**Table 5** (regression #1) shows the results of multiple regression analysis with TS_ phenome, phenome, or SB as dependent variables and the ACEs and BFI subdomain scores as explanatory variables, while allowing for the effects of background variables such as age, sex, education, etc. We found that 30.3% of the TS_phenome was explained by the combined effects of PC_neglect, PC_sexabuse, PC_abuse (all positively associated) and age (inversely associated). **Figure 1** shows the partial regression of TS_phenome on PC_neglect. Regression #2 shows that 70.2% of the variance in the phenome score was explained by the regression on neuroticism, PC_neglect (both positively) and age (inversely associated). **Figure 2** shows the partial regression of the phenome score on neuroticism. Introduction of dysthymia (regression #3) improved the prediction by 3.1%. We found that 36.1% in the SB score (regression #4) was explained by PC_4BFI and PC_sexabuse. Regression #5 shows that neuroticism is predicted by the three ACE domains (all positively associated) and age (inversely associated). **Figure 3** shows the partial regression of neuroticism on neglect. The 4 other BFI subdomain scores were differently predicted by the three ACE PC scores, with or without age or sex, albeit with a much lower effect size as compared with neuroticism.

**Table 5.** Results of multiple regression analyses with phenome data and personality domains as dependent variables and adverse childhood experiences as input variables

| Dependent variables | Explanatory variables | Parameter estimates + statistics |  |  | Model statistics + effect sizes |  |  |  |
| --- | --- | --- | --- | --- | --- | --- | --- | --- |
| | | $\beta$ | t | p | F<sub>model</sub> | df | p | R<sup>2</sup> |
| #1 TS_phenome | **Model** |  |  |  | **13.88** | **4/128** | **<0.001** | **0.303** |
|  | PC_neglect | 0.297 | 3.54 | <0.001 |  |  |  |  |
|  | Age | -0.263 | -3.55 | <0.001 |  |  |  |  |
|  | PC_sexabuse | 0.185 | 2.46 | 0.015 |  |  |  |  |
|  | PC_abuse | 0.169 | 2.00 | 0.048 |  |  |  |  |
| #2. Phenome | **Model** |  |  |  | **101.08** | **3/129** | **<0.001** | **0.702** |
|  | BFI_neuroticism | 0.702 | 12.90 | <0.001 |  |  |  |  |
|  | PC_neglect | 0.197 | 3.71 | <0.001 |  |  |  |  |
|  | Age | -0.131 | -2.65 | 0.009 |  |  |  |  |
| #3. Phenome | **Model** |  |  |  | **88.71** | **4/128** | **<0.001** | **0.735** |
|  | BFI_neuroticism | 0.615 | 11.02 | <0.001 |  |  |  |  |
|  | Dysthymia | 0.203 | 4.01 | <0.001 |  |  |  |  |
|  | PC_neglect | 0.197 | 3.92 | <0.001 |  |  |  |  |
|  | Age | -0.123 | -2.63 | 0.010 |  |  |  |  |
| #4. Current SB | **Model** |  |  |  | **36.68** | **2/130** | **<0.001** | **0.361** |
|  | PC_BFI | 0.493 | 6.91 | <0.001 |  |  |  |  |
|  | PC_sexabuse | 0.264 | 3.71 | <0.001 |  |  |  |  |
| #5. Neuroticism | **Model** |  |  |  | **14.12** | **4/128** | **<0.001** | **0.306** |
|  | PC_neglect | 0.279 | 3.34 | 0.001 |  |  |  |  |
|  | PC_abuse | 0.227 | 2.69 | 0.008 |  |  |  |  |
|  | PC_sexabuse | 0.199 | 2.64 | 0.009 |  |  |  |  |
|  | Age | -0.195 | -2.64 | 0.009 |  |  |  |  |
| #6. Extraversion | **Model** |  |  |  | **11.83** | **1/131** | **<0.001** | **0.166** |
|  | PC_neglect | -0.298 | -3.71 | <0.001 |  |  |  |  |
|  | Age | 0.209 | 2.60 | 0.010 |  |  |  |  |
|  | Sex | -0.186 | -2.21 | 0.022 |  |  |  |  |
| #7. Agreeableness | **Model** |  |  |  | **10.64** | **1/131** | **0.001** | **0.075** |
|  | PC_neglect | -0.274 | -3.26 | 0.001 |  |  |  |  |
| #8. Conscientiousness | **Model** |  |  |  | **10.46** | **2/130** | **<0.001** | **0.138** |
|  | Age | 0.276 | 3.39 | <0.001 |  |  |  |  |
|  | PC_abuse | -0.235 | -2.88 | 0.005 |  |  |  |  |
| #9 Openness | **Model** |  |  |  | **6.42** | **2/130** | **0.002** | **0.090** |
|  | Education | 0.246 | 2.92 | 0.004 |  |  |  |  |
|  | PC_sexabuse | 0.197 | 2.34 | 0.021 |  |  |  |  |

TS\_phenome: trait-state-Phenome; PC: principal component, BFI: Big Five Inventory; SB: suicidal behaviors

**Figure 1.**
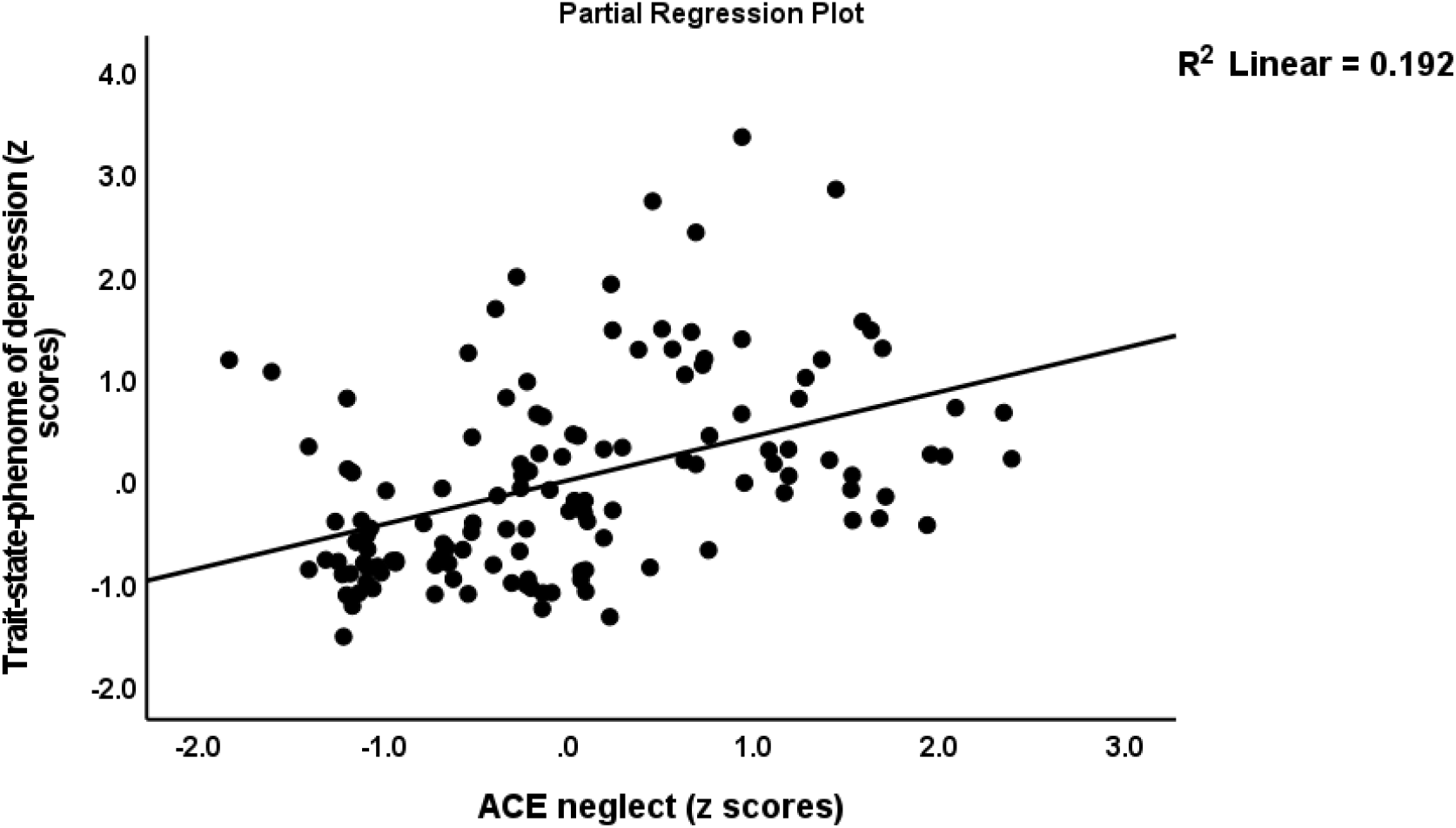
Partial regression of the trait-state phenome on neglect adverse childhood experiences (ACE)

**Figure 2.**
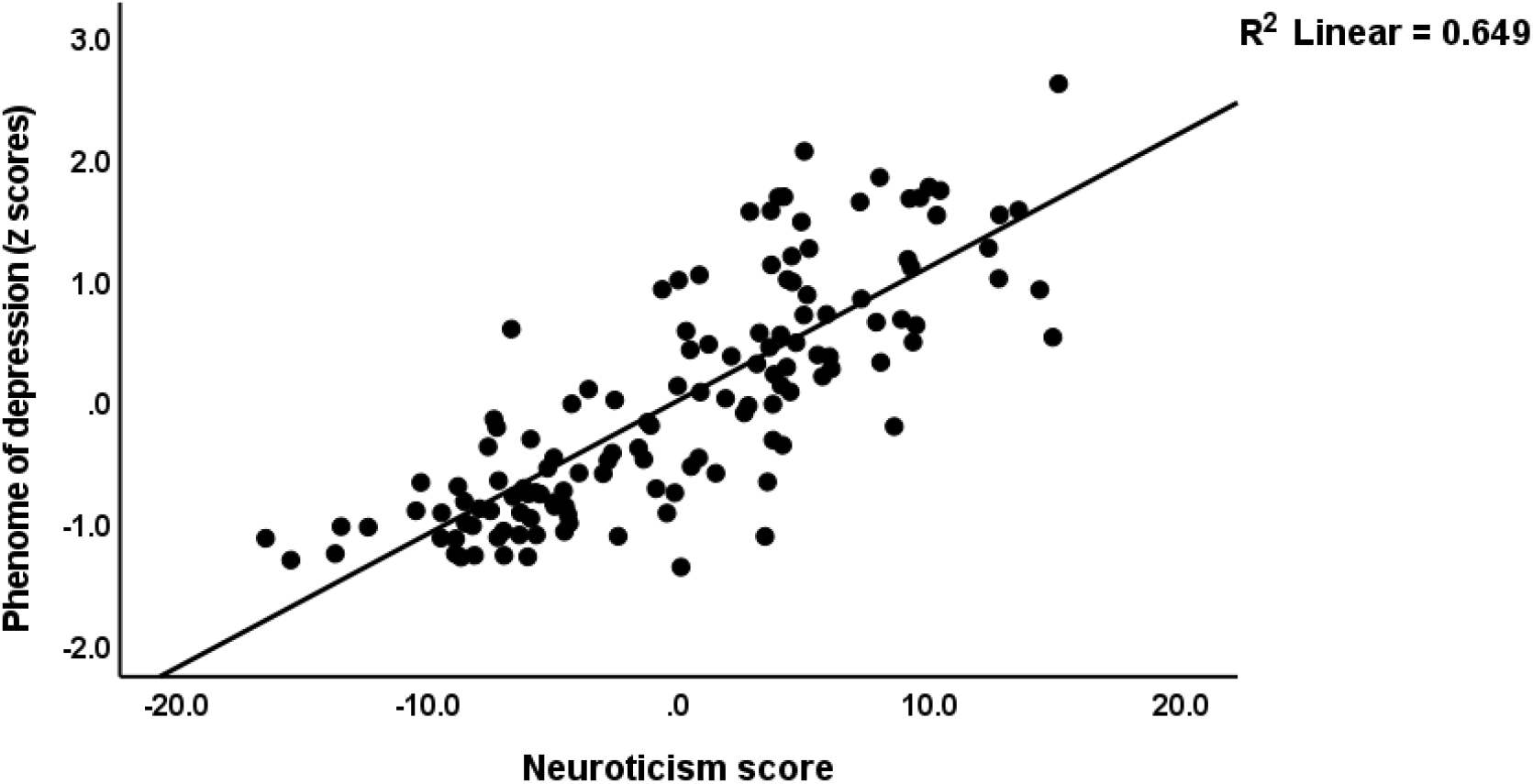
Partial regression of the phenome of depression on neuroticism

**Figure 3.**
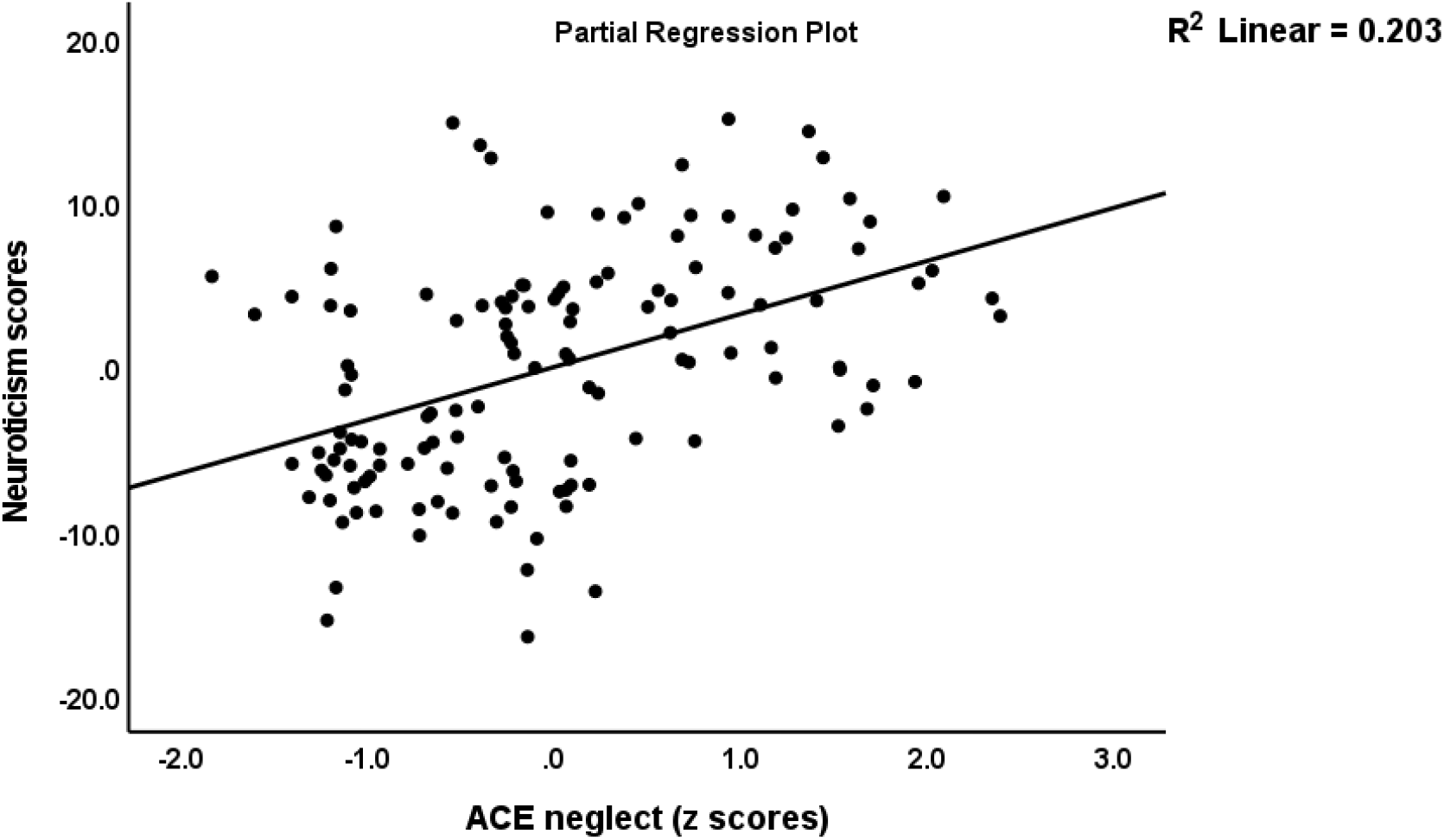
Partial regression of neuroticism on neglect adverse childhood experiences (ACE)

#### Results of PLS analysis

Figure 4. shows the results of PLS analysis after feature and path selection. The phenome was conceived of as a latent vector derived from HAMD, BDI, STAI and SB scores. Predictors were the 5 BFI subdomain scores, the LVs constructed using neglect, abuse and sexabuse ACE scores, age, sex, and a first-degree FHIS of mood disorders + SB and FHIS of SUD. LV_neglect was conceptualized as the first factor extracted from 6 ACE items, and LV_abuse as the first factor extracted from 7 ACE items. We considered that the BFI domains could mediate the effects of ACEs on the phenome. Moreover, we examined moderation models whereby ACE moderate the effects of BFI domains. The three latent vectors constructed here showed adequate construct validity and convergence: a) all AVEs were > 0.50, namely for LV_phenome: 0.898, ACEneglect: 0.739 and ACEabuse: 0.573; b) the Cronbach’s alpha for the three LV constructs were 0.962, 0.929 and 0.877, respectively; the composite reliabilities 0.972, 0.944 and 0.903, respectively; c) the SRMR is 0.046; d) PLSPredict shows that the Q^2^ predict values are positive (suggesting that the constructed model outperforms the most naïve benchmark); and e) the Heterotrait-Monotrait ratio showed that discriminatory validity is established. We found that 74.3% of the variance in the phenome score was explained by the regression on LV_neglect, neuroticism (both positively), and age (inversely). Moreover, there was a significant interaction (moderation) between LV_neglect and neuroticism, yielding a positive effect on the phenome. We found that 29.4% of neuroticism was explained by both LV_neglect and LV_abuse. LV_neglect also affected agreeableness and extraversion, whereas LV_abuse also impacted conscientiousness, although only neuroticism yielded a direct effect on the phenome. There were specific indirect effects of LV_abuse (t=3.79, p<0.001) and LV_neglect (t=3.57, p<0.001) on the phenome which were mediated by neuroticism. There were significant total effects of a family history of mood disorders + SBs (t=3.09, p=0.002) on the phenome which were mediated by the paths from LV_neglect and LV_abuse to neuroticism. PLS multigroup analysis and premutation analyses showed that there were no significant differences among men and women and among those with and without metabolic syndrome in any of the model features.

### Discussion

#### Neuroticism, depression and suicidal behaviors

The first major finding of this study is that neuroticism is significantly higher and extraversion and agreeableness are significantly lower in MDD patients compared to controls, whereas there were no significant differences in conscientiousness and openness. Previous research also demonstrated that BFI-measured personality traits are associated with major depression, depressive phenotypes, and depression severity (Kotov et al., 2010, Allen et al., 2017, Lester, 2021, Koorevaar et al., 2017). For instance, according to Lester (Lester, 2021), the five BFI personality scores were correlated with depression severity scores and accounted for 36% of the variance. Neuroticism, agreeableness, conscientiousness, and extraversion were associated with atypical or melancholic major depression in older adults (Koorevaar et al., 2017). Jourdy and Petot (Jourdy and Petot, 2017) discovered significant linear associations between quantitative depression severity scores and neuroticism and conscientiousness, but not extraversion, extroversion, or agreeableness. A quantitative review of 175 studies (1980–2007) on the Big Three Inventory and BFI revealed that depressed patients had higher neuroticism and lower conscientiousness scores than controls, while unipolar depression was associated with higher neuroticism and lower extraversion and conscientiousness scores (Kotov et al., 2010). A recent meta-analysis (2022) comprising 243 studies that examined the associations between BFI traits and depression found a positive correlation between depression and neuroticism scores and a significant inverse correlation with conscientiousness, extraversion, and openness (Chavoshi, 2022).

**Figure 4.**
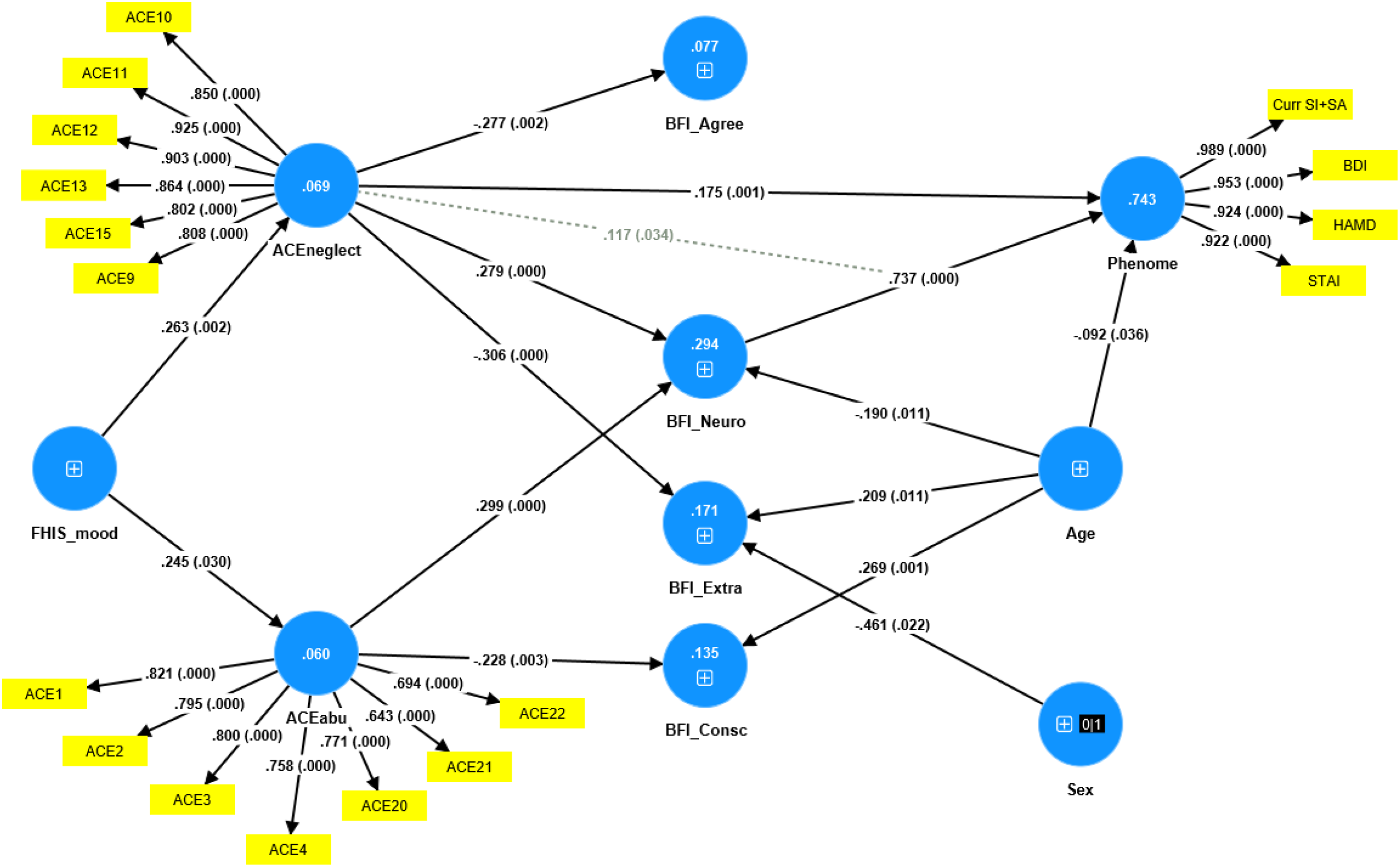
Results of PLS analysis. Shown are the significant paths. The phenome, neglect adverse childhood experiences (ACEneglect) and ACEabuse were entered as latent vectors extracted from their manifestations. The other variables were entered as single indicators (denoted as +); sex was entered as a binary variable (1=male, 0=female). Curr SI+SA: current suicidal ideation + attempts; BDI: Beck Depression Inventory; HAMD: Hamilton Depression Rating Scale; STAI: State and Trait Anxiety Inventory; ACE items: see supplementary file, Table 2 for an explanation; FHISmood: a family history of mood disorders and suicidal attempts. BFI: Big Five Inventory; Agree: agreeableness, Neuro: neuroticism, Extra: extraversion, Consc: conscientiousness. Shown are the path coefficients (with exact p value) of the inner model, and loadings (with p values) of the outer model; figures in the blue circles: explained variance.

In the stress-diathesis model of depression, neuroticism is considered one of the important diathesis factors that increases the risk of depression and depression severity (Chavoshi, 2022). As a result, it is possible to conclude that elevated neuroticism in conjunction with decreased extraversion and agreeableness (our study) and decreased openness and conscientiousness (see previous studies) increases the risk of depression and suicidal behaviors. However, we discovered that one valid and reliable factor could be extracted from neuroticism (negatively loaded), extraversion, agreeableness, and conscientiousness (all positively loaded), and that this factor demonstrated adequate convergent and reliability validity. Consequently, these four domains are manifestations of a common core, which can be termed “the NACE dimension” (all first letters of the 4 personality traits). This may appear to contradict the 5-factor structure of the BFI, which was developed using factor analysis (Srivastava, 2016). These factors were derived from Western study samples, whereas our research involved Thai participants. While population differences may play a role, the most likely explanation for our one-factor model (the NACE dimension) is that the BFI assessment in depression biases trait measurements toward a more depressive phenotype, characterized by changes in motivation, autobiographical memories, attitude, emotion, self-concepts, and social roles.

In this regard, Allen et al. (Allen et al., 2017) proposed that depressive tendencies are determined by the “best two out of three” interaction pattern between a high-risk (withdrawal) and protective (industriousness and enthusiasm) domains. Notwithstanding, our findings indicate that a common trait (NACE-dimension) is associated with the severity of the depressive state and that neuroticism has a greater influence on the phenome of acute depression than PC_NACE. Therefore, neuroticism may be considered the “worst of five traits.” These findings support those of Kotov et al. (Kotov et al., 2010) who concluded that neuroticism is the best predictor of psychopathology.

Another question is whether the depressive state (the phenome of depression) can influence BFI characteristics. In this regard, small effects of depression and anxiety in the acute state on neuroticism were observed (Karsten et al., 2012). The depressive state affected extraversion and conscientiousness, but not agreeableness and openness (Karsten et al., 2012). There is now evidence that personality traits are not fixed, genetically determined traits, but can change over time in response to environmental factors, such as the effects of depression (McCrae and Costa Jr, 1999, Srivastava et al., 2003, Roberts et al., 2006, Costa et al., 2005, Karsten et al., 2012). Similarly, extraversion may decline during depressive episodes and even increase permanently thereafter (Griens et al., 2002, Kendler et al., 1993). During depressive phases, however, agreeableness, conscientiousness, and openness were found to be more stable (Malouff et al., 2005). Lesser known are the effects of anxiety on personality traits (Karsten et al., 2012).

In the present study, however, we discovered that neuroticism accounted for 64.9% of the variance in the severity of depression and that one replicable and validated factor could be extracted from severity of depression, anxiety, suicidal behaviors, and neuroticism. This suggests that neuroticism (assessed as a trait), severity of depression/anxiety (assessed as a state over the last week), and suicidal behaviors (assessed as a state over the last month) are manifestations of a common core that may be termed the “trait-state-(TS) phenome of depression.” In addition, one factor could be extracted from the NACE trait dimension, the depression phenome (a PC extracted from depression, anxiety, and suicidal behaviors), and a positive lifetime history of dysthymia and anxiety disorders. In this respect, dysthymia, GAD, PTSD, panic disorder, agoraphobia, social phobia, and OCD were strongly associated with neuroticism, whereas dysthymia and certain anxiety disorders were also associated with extraversion and conscientiousness (both inversely) (Kotov et al., 2010).

In the 1970s and 1980s, some scientists conceptualized “neurotic depression” as depression in patients with pre-existing characteristics, such as emotional instability, anxiety, maladaptive coping, internalization, fixation on negative memories, maladaptive processing, low self-esteem, etc (Klerman et al., 1979, Winokur et al., 1987, Nierenberg and Ellard, 2015, Chamberlain, 2023). Other researchers have hypothesized that emotional dysregulation and maladaptive coping could mediate the relationship between neuroticism and “neurotic depression” (Guo et al., 2003, Yoon et al., 2013). As a result, neurotic depression has been proposed as yet another subtype of depression (Chamberlain, 2023), adding to the already absurdly long list of depressive subtypes (Maes et al., 2022b, Maes and Stoyanov, 2022). In spite of this, the fact that a single factor can be extracted from personality (a trait), lifetime diagnosis of dysthymia and anxiety disorders (traits), and the acute phenome of depression (state) suggests that these trait and state facets are manifestations of a common core and share a common pathophysiology. Moreover, all components of neuroticism are potential symptoms of depression (is blue, not relaxed, tense, worries, unstable, moody, not calm, nervous). In addition, a portion of the extraversion and agreeableness domain items (which in our study are both decreased in depressed patients) may appear as symptoms of depression, such as less talkative, less energy, less enthusiasm, be quiet, less assertive, inhibited, less sociable (from the extraversion domain), and having quarrels with others, not trusting others, being aloof, and having less cooperation with others (items belonging to the agreeableness trait). Therefore, it is more appropriate to conclude that neuroticism is a “forme fruste” of major depressive disorder, either as a prodrome (pre-symptoms), subclinical symptoms, or residual symptoms following an acute episode.

#### ACEs, BFI domains and the phenome of depression

The second major finding of this study is that a large proportion of the variance in neuroticism (29.4%) is explained by the effects of ACEs, specifically the combined effects of neglect and abuse (both physical and emotional). In addition, higher neglect scores predicted lower agreeableness and extraversion, while abuse predicted lower conscientiousness. Furthermore, neuroticism partially mediated the effects of neglect on the phenome, and there was an additional moderating (or interaction) effect, namely an interaction between neglect and neuroticism. The effects of abuse on the phenome were totally mediated by neuroticism. These findings expand upon those of prior studies indicating that early-life trauma may contribute to neuroticism (Barlow et al., 2014, Lahey, 2009, Winokur et al., 1987). There is now overwhelming evidence that ACEs are predictors of major depression, depression severity, and suicidal behaviors (Maes et al., 2018, Maes et al., 2019, Maes et al., 2021, Maes et al., 2022b, Maes et al., 2023, Maes et al., 2021).

Moreover, the common trait-state phenome of depression (thus the first PC extracted from neuroticism and the phenome of depression including suicidal behaviors) is predicted to a significant degree (approximately 30%) by the combined effects of neglect, abuse, and sexual abuse. These results suggest that a common denominator (ACEs) predicts trait (low grade depression or neuroticism) and state (the acute phenome of depression) phenomena, which are both indicators of the “chronic disorder” major depression. Phrased differently, these findings also indicate that neuroticism is a forme fruste of a major depressive episode.

#### Explanatory mechanism underpinning the effects of ACEs on depression

Recent research indicates that ACEs may influence the microimmuneoxysome (changes in the microbiome and immune and oxidative pathways), resulting in increased neurotoxicity and, consequently, depressive symptoms (Maes et al., 2023). First, increased ACEs are associated with a particular compositional gut dysbiosis enterotype, which mediates the effects of ACEs on the severity of the depression phenotype (Maes et al., 2023). This is significant because depression is associated with an increase in bacterial translocation caused by a leaky gut (Maes et al., 2008). The latter is fueled by the depression-associated enterotype, which promotes abnormalities in epithelial cells, the gut barrier, leaky gut, and bacterial translocation, resulting in increased plasma LPS concentrations that stimulate the Toll-like receptor (TLR)4 complex and thus inflammatory and oxidative pathways (Lucas and Maes, 2013, Maes et al., 2023). Second, ACEs may lead to decreased antioxidant levels, such as paraoxonase 1 (PON 1) and high-density lipoprotein cholesterol, as well as activated nitro-oxidative stress pathways (Maes et al., 2019, Maes et al., 2022b), which, in combination with elevated plasma LPS, may result in a breakdown of the blood-brain barrier and increased neurotoxicity (Maes et al., 2023). Thirdly, ACEs induce immune sensitization with increased activation of the cytokine and growth factor networks following ex vivo administration of LPS and mitogens to whole blood (Maes et al., 2022b). This suggests that ACEs sensitize cytokine/growth factor networks, which are activated upon stimulation with stressors such as LPS (e.g., as a result of leaky gut or gut dysbiosis), bacterial or viral infections, and psychosocial stressors (Maes et al., 1999, Maes et al., 2022b). The latter causes activation of T helper 1 and M1 macrophages as well as elevated levels of interferon-γ, interleukin-6, and tumor necrosis factor-α, which are all associated with stress-induced anxiety (Maes et al., 1999). Consequently, it is tempting to hypothesize that ACE-sensitized microimmuneoxysome pathways result in a low-grade depression trait (neuroticism or prodrome or subclinical symptoms) or major depressive episodes in response to a variety of stressors. Biomarker research should examine whether neuroticism and major depression share the same biomarkers, and if so, this would further indicate that neuroticism is, indeed, a forme fruste of depression.

### Limitations

This study would have been more interesting if we had also determined the cytokine and growth factor network and oxidative stress pathways. It could be argued that the sample size is relatively small. However, an *a priori* calculation of the sample size showed that a sample size of at least n=70 is required to achieve a power of 0.80 at p=0.05 (two tailed). Moreover, the regressions (including the PLS analysis) of the phenome on the neuroticism and 2 other indicators revealed that, given the study sample of 133 participants (tested at alpha=0.05 and 3), the actual power was 1.

### Conclusions

Neuroticism scores are significantly higher in MDD than in controls and accounted for 64.9% of the variance in the depression phenome. Neuroticism is a much better predictor of the phenome than extraversion and agreeableness, whilst openness and conscientiousness do not have any significant effect. One latent vector could be extracted from neuroticism, dysthymia, lifetime anxiety disorders (trait features) and the phenome of depression (a state features assessed over the last month or week). Moreover, physical and emotional neglect, physical, emotional and sexual abuse account for about 30% of the variance in this latent construct. PLS analysis shows that the effects of neglect on the phenome are partially mediated by neuroticism, whereas the effects of abuse were entirely mediated by neuroticism. Neuroticism and major depressive disorder are manifestations of the same latent core, which is in part caused by ACEs. Neuroticism is a less severe manifestation of major depressive disorder and the latter is a magnified manifestation of “neuroticism”. Therefore, the latter should be better described as a forme fruste of major depression, rather than a personality trait. Future research should examine whether the microimmuneoxysome biomarkers of major depression are also biomarkers of neuroticism which would reinforce the idea that neuroticism is a preclinical stage of major depression. Most important is to examine whether neuroticism is associated with the same sensitized cytokine/growth factors networks as MDD and whether reactivation of these pathways through negative life events, infections, or inflammatory stressors is associated with a transitions form neuroticism to a full blown major depressive episode. Both neuroticism and major depression are manifestations of the same latent core, whereby the former is a forme fruste of major depression, and the latter a magnified manifestation.

## Data Availability

All data produced in the present study are available upon reasonable request to the authors.

## Author’s contributions

Conceptualization and study design: MM and JK; first draft writing: KJ and MM; editing: all authors; recruitment of patients: JK. All authors approved the final version of the manuscript.

## Ethics approval and consent to participate

The research project (#445/63) was approved by the Institutional Review Board of Chulalongkorn University’s institutional ethics board. All patients and controls gave written informed consent priot to participation in the study.

## Data Availability Statement

The dataset generated during and/or analyzed during the current study will be available from the corresponding author upon reasonable request and once the dataset has been fully exploited by the authors.

