## Supplemental Table 1 and 2 for "The effects of adverse childhood experiences on depression and suicidal behaviors are partially mediated by neuroticism, a forme fruste of major depression"

**ELECTRONIC SUPLEMENTARY FILE (ESF)**

**ESF Table 1**. Adverse childhood experiences (ACEs) Questionnaire

| **ACE** | **Questions** |
|---|---|
| ACE 1 | You have been reviled, insulted and despised. |
| ACE2 | You have encountered some behaviors that make you afraid of getting hurt. |
| ACE3 | You have been violently pushed, grabbed, slapped, yanked, or hurled. |
| ACE4 | You have been hit so violently that you left a mark or were injured. |
| ACE5 | You have been sexually touched or fondled on your body. |
| ACE6 | You have experienced someone letting you touch their bodies in a sexual way. |
| ACE7 | You have experienced someone trying to have sex with you in different ways(oral,anal,vagina). |
| ACE8 | You have been forced to have sex in different ways (for example, oral sex, anal sex, and vagina sex). |
| ACE9 | Certain family members make you feel important and special. |
| ACE10 | You feel that you are beloved. |
| ACE11 | Your family members care about and love each other. |
| ACE12 | Your family members have deep connections. |
| ACE13 | Your family can be your strength and support. |
| ACE14 | You eat an insufficient diet. |
| ACE15 | You are aware that you have been shielded and take precautions. |
| ACE16 | Your parents are alcohol and drug addicted, so they cannot take care of the family. |
| ACE17 | You need to wear dirty clothes. |
| ACE18 | When you need to see a therapist, someone will bring you. |
| ACE19 | Your mother or stepmother have been violently pushed, grabbed, slapped, yanked, or hurled |
| ACE20 | Your mother or stepmother have been kicked, bit, or punched with fists or solid objects. |
| ACE21 | Your mother or stepmother have been repeatedly beaten, even after a few minutes. |
| ACE22 | Your mother or stepmother have been threatened or wounded with a knife or a gun. |
| ACE23 | You live with someone who is a heavy drinker or an alcoholic, which causes troubles. |
| ACE24 | You live with someone who is on drugs. |
| ACE25 | You have family members who have depression or mental disorders. |
| ACE26 | You have family members who have attempted or completed suicide. |
| ACE27 | Your parents have separated or divorced. |
| ACE28 | Your family members have been in prison. |

**ESF, Table 2**. Columbia Suicide Severity Rating Scale (C-SSRS)

| | |
|---|---|
| Current suicidal ideation 7 C-SSRS items, namely | |
| C-SSRS11 | Wish of dead |
| C-SSRS12 | Non-specific active suicidal thoughts |
| C-SSRS13 | Active Suicidal Ideation with Any Methods (Not Plan) without Intent to Act |
| C-SSRS15 | Active Suicidal Ideation with Specifie Plan and Intent |
| C-SSRS16 | How many times have you had these thoughts? |
| CSSRS17 | When you have the thoughts, how long do they last? |
| Current suicidal attempt 5 C-SSRS items, namely | |
| C-SSRS30 | Actual Attempt |
| C-SSRS31 | Have you made a suicide attempt? (Total of Attempts) |
| C-SSRS32 | Has subject engaged in Non-Suicidal Self-Injurious Behavior? |
| C-SSRS33 | Interrupted Attempt |
| C-SSRS34 | Has there been a time when you started to do something to end your life but someone or something stopped you before you actually did anything? (Total of interrupted) |
